# What is the relationship between raising the minimum legal sales age of tobacco above 20 and cigarette smoking? A systematic review

**DOI:** 10.1101/2023.10.18.23296747

**Authors:** Nathan Davies, Ilze Bogdanovica, Shaun McGill, Rachael L Murray

**Author notes:** **Corresponding author**, 0115 844 1514.

## Abstract

**Objectives:** To determine whether raising the minimum legal sales age of tobacco (MLSA) to 20 or above is associated with reduced prevalence of smoking compared to an MLSA set at 18 or below through systematic review.

**Data sources:** Following a pre-registered protocol on PROSPERO (ref: CRD42022347604), six databases of peer-reviewed journals were searched from January 2015 to September 2023. Backwards and forwards reference searching was conducted.

**Study selection:** Studies that assessed the association between MLSAs of 20 and above with cigarette smoking or cigarette sales for children and young people aged 11-20. Assessments on e-cigarettes were excluded.

**Data extraction:** Pairs of reviewers independently extracted study data and used ROBINS-I to assess risk of bias.

**Data synthesis:** Narrative methods were used to synthesise findings. 19 studies were reviewed, from which 26 effect estimates were extracted. All studies evaluated Tobacco 21 laws in the United States. Just under half of estimates found a statistically significant association with reduced current cigarette smoking or sales, just over half found no statistically significant association, and one estimate found an association with increased cigarette smoking. The positive association appeared to be stronger for older age groups, those from a Hispanic/Latinx background and those with lower education. The degree of study bias was variable.

**Conclusions:** There is evidence that raising the MLSA for tobacco to 21 reduces cigarette sales and current cigarette smoking amongst those aged 11-20 and has potential to reduce health inequalities. Further research beyond the United States would support generalisability to other settings.

## INTRODUCTION

Globally, over 80% of tobacco smokers start smoking aged 15-24.[1] In 2019, 155 million in this age group were regular tobacco smokers.[1] Preventing initiation of and regular tobacco use in this age group, in which individuals are markedly susceptible to addiction[2] is critical to prevent future smoking harm. Minimum legal sales age laws (MLSA), which prohibit retailers and vendors from selling tobacco products to those under a certain age, are one policy option for reducing access to tobacco products.

There is good evidence that raising the MLSA to 18 is associated with reduced smoking rates in the target population in England [3–5] and reduced commercial tobacco purchases in Finland, [6] although a study on raised European MLSAs did not find an association with smoking prevalence.[7] Given the evidence base for MLSAs, and the uniquely harmful properties of tobacco, there has been renewed global interest in increasing MLSAs beyond 18. [8–12] Article 16 of the World Health Organization Framework Convention for Tobacco Control compels signatories to prohibit the sale of tobacco products to minors but does not specify an exact age limit.[13]

In the 21^st^ century, following the lead of Needham, Massachusetts, local areas, cities and states across the US began to introduce Tobacco 21 (T21) laws [14] culminating in a national law being passed in 2019.[15] Several other countries, including Ethiopia, Honduras, Japan, Kazakhstan, Mongolia, Philippines, Singapore, Sri Lanka, Thailand, Turkmenistan and Uganda have been reported to introduce a MLSA of at least 20 in recent years.[16] New Zealand recently introduced a smoke-free generation law intended to ban sales of tobacco to anyone born after 2009 [17] and the UK government is consulting on a similar policy.[18] As policymakers across the globe continue to consider raising MLSA laws as an important component of strategies to reduce tobacco harm, understanding the effectiveness of such policies is of great importance. The main objective of this systematic review was to determine if raising the legal age of sale of tobacco to 20 or above is associated with reduced prevalence of smoking amongst those aged 11-20, compared to a legal age of tobacco set at 18 or below.

## METHODS

This systematic review and meta-analysis was conducted in line with a pre-registered protocol on PROSPERO (ref: CRD42022347604) and the Preferred Reporting Items for Systematic Review and Meta-Analyses (PRISMA) reporting guidelines (Supplementary File 1).[19]

### Selection criteria

Studies were eligible if they reported the effect on cigarette use of raising the MLSA to 20 or above. They were eligible if the study population included children and young people aged 11-25, or if data restricted to this age group could be extracted from the broader study. We excluded studies where a MLSA of 20 or above was introduced where no prior age-of-sale limit previously existed. We excluded qualitative studies and studies that purely reported estimates relating to e-cigarettes. There were no geographical restrictions.

### Search strategy and study selection

We searched the electronic databases Embase through OVID, MEDLINE through PubMed, PyscINFO through Ovid, ProQUEST Public Health, ProQUESTION Dissertations and Theses, and CINAHL through Ebscohost, for studies published from 1 January 2015 (the year in which the first study evaluating the local T21 law in Needham, Massachusetts, was published) to 15 September 2023. A full list of search terms is provided in Supplementary File 2. No restrictions were in place for the observational period or language. Records were extracted into Rayyan[20] and de-deduplicated. Two of three reviewers (ND, IB, RM) screened titles and abstracts, and subsequently full texts to identify eligible studies. ND hand searched reference lists of identified studies to identify any additional studies. Conflict was settled by discussion or by adjudication from the third screener.

### Data extraction and quality assessment

A standard data extraction form was piloted and used to record details for each eligible study by two reviewers (ND and IB). Two of three reviewers (ND, RM, SM) independently assessed quality using the Risk of Bias in Non-randomized Studies – of Interventions (ROBINS-I) assessment tool [21] with conflict resolved by discussion and adjudication from a third reviewer where necessary. Studies were given a risk of bias score for 7 domains between “low” and “critical” and given an overall risk of bias score.

### Data synthesis

We planned to conduct a meta-analysis and sensitivity analyses. However, the data was not found to be suitable for meta-analysis. Even after approaching authors for further information, many studies eligible for inclusion used measures of effect that could not be harmonised, including several studies with moderate risk of bias. Furthermore, studies included very heterogeneous population groups, comparators, and outcome measures.

Instead, narrative synthesis was conducted, following the process of developing a preliminary synthesis, exploring relationships within and between studies through visual tabulation of intervention type, population characteristics, measures of effect, and whether a statistically significant association was found between current smoking or cigarette sales and T21. The robustness of the synthesis was assessed by considering the quality of included studies.[22] As no primary patient-level data was used, ethical approval was not required.

## RESULTS

### Overview of included studies

Database searches identified 3,180 papers, of which 2,485 were unique papers. 31 papers remained after title and abstract screening. 11 were judged ineligible in the full-text review; six papers had no outcome data, two papers did not relate to age-of-sale policy, two papers related to age-of-sale policies restricting sales to those under 20 only, and one was not peer-reviewed. One paper was identified during citation searching, resulting in 19 papers being included. (Figure 1).

**Figure 1:**
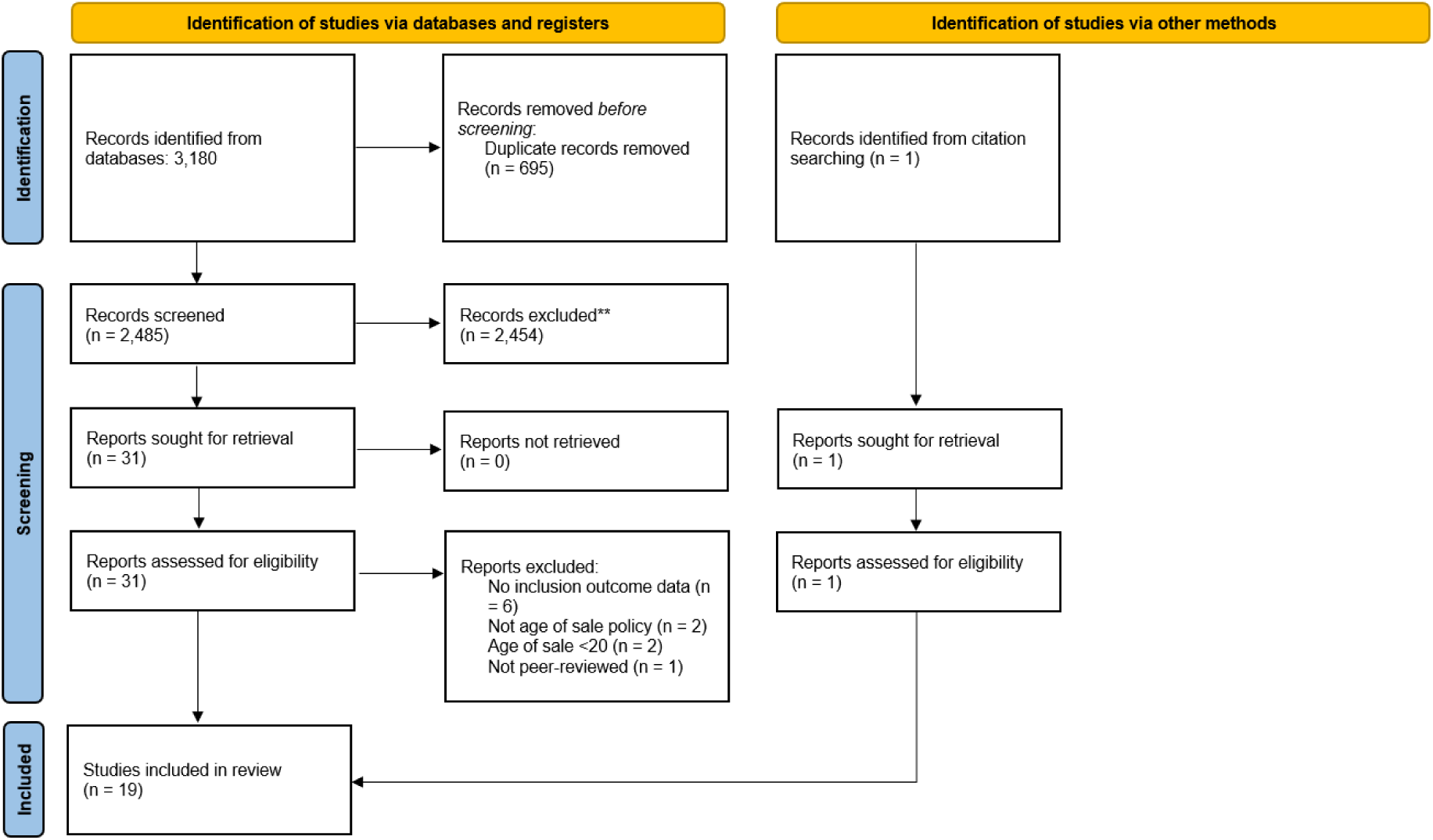
PRISMA flow diagram

26 estimates of association of the policy with current smoking or cigarette sales were extracted from the 19 remaining studies.[23–41] (Table 1). Full data extraction is available in Supplementary File 3.

**Table 1:** Summary of studies.

| Study author | Design | Data collection | Intervention type | Intervention group | Age | Comparator | Outcome | Direction of association | Risk of bias |
| --- | --- | --- | --- | --- | --- | --- | --- | --- | --- |
| **Friedman 2019**[23] | One-off cross-sectional | Nov 2016 to May 2017 | Large local area and state laws | 18-20 in T21 area who has ever tried cigarette (US) (n=911) | 18+ | 21-22 in Tobacco-21 area who has ever tried a cigarette (US) (n=958) | Current cigarette smoking: AOR 0.6 (CI 0.39, 0.92)^ | Favours intervention | Serious |
| **Macinko 2018**[24] | Repeated cross-sectional | 2008-2016 | New York City T21 law | 7-12 graders living in New York City (n=76,668) | U18 | 7-12 graders year olds living in New York State (n= 76,668) | Current cigarette smoking: APR 1.25 (0.88, 1.76) | Not significant | NI |
| **Macinko 2018**[24] | Repeated cross-sectional | 2007-2015 | New York City T21 law | 9-12 graders living in New York City (n= 71,214*) | U18 | 9-12 graders year olds living in 4 cities in Florida (n= 71,214*) | Current smoking (cigarette only): APR 1.40 (1.10, 1.80)^ | Favours control | Serious |
| **Garcia-Ramirez 2022**[25] | Repeated cross-sectional | 2013-2019 | California state T21 Law | 7, 9, and 11 graders in public school in California post-policy (n=2,229,401*) | U18 | 7, 9, and 11 graders in public school in California pre-policy (n=2,229,401*) | Current cigarette smoking: AOR 0.98 (0.94, 1.03) | Not significant | Moderate |
| **Friedman 2020**[26] | Repeated cross-sectional | 2011-2016 | Local laws | 18-20 living in local T21 area (n= 4,813) | 18+ | 18-20 not in T21 area (n = 20,253) | Current cigarette smoking: -0.0306 absolute risk reduction (CI -0.0548 to -0.0063)^ | Favours intervention | Moderate |
| **Agaku 2022**[27] | One-off cross-sectional | 2019 | State laws | 18-20 living in state T21 area (n=10,146*) | 18+ | 18-20 not living in state T21 area (n=10,146*) | Current cigarette smoking: APR 0.58 (0.39, 0.74)^ | Favours intervention | Serious |
| **Agaku 2022**[27] | One-off cross-sectional | 2019 | State laws | Grades 9-12 living in state T21 area (n=182,491*) | U18 | Grades 9-12 living in state T21 area (n=182,491*) | Current cigarette smoking: APR 0.70 (0.52-0.93)^ | Favours intervention | Serious |
| **Dove 2021**[28] | Repeated | 2012-2019 | California state | 18-20 year olds in California (n | 18+ | 18-23 year olds in 8 referent | Current cigarette smoking: 1.01 (0.76, | Not significant | Serious |
|  | cross-sectional |  | T21 law | = 3050) |  | states (n = 12,813) | 1.34) |  |  |
| **Colston 2022**[29] | Repeated cross-sectional | 2013-2019 | Local, county and state laws | 8 graders in T21 area (n=92,922*) | U18 | 8 graders without T21 coverage (n=92,922*) | Current cigarette smoking: ARR 0.91 (0.69, 1.20) | Not significant | Moderate |
| **Colston 2022**[29] | Repeated cross-sectional | 2013-2019 | Local, county and state laws | 10 graders in T21 area (n=88,628*) | U18 | 10 graders without T21 coverage (n=88,628*) | Current cigarette smoking: ARR 0.96 (0.75, 1.23) | Not significant | Moderate |
| **Colston 2022**[29] | Repeated cross-sectional | 2013-2019 | Local, county and state laws | 12 graders in T21 area (n=81,082*) | U18 | 12 graders without T21 coverage (n=81,082*) | Current cigarette smoking: ARR 0.74 (0.60 - 0.91)^ | Favours intervention | Moderate |
| **Roberts 2022**[30] | Cohort study | 2016 and 2018 | Columbus, Ohio T21 law | First-year students pre-T21 in Columbus (n=529) | 18+ | First-year students post-T21 in Columbus (n = 611) | Current cigarette smoking: 4.1% (interv.) 6.6% (control) | Not significant | Critical |
| **Grube 2021**[31] | Repeated cross-sectional | 2010 - 2018 | California state T21 Law | 7, 9, and 11 graders in public school in California post-policy (n=2,956,054*) | U18 | 7, 9, and 11 graders in public school in California pre-policy (n=2,956,054*) | Current cigarette smoking: AOR 0.99 (0.97, 1.01) | Not significant | NI |
| **Glover-Kudon 2021**[32] | Longitudinal cigarette sales data | 2012 - 2017 | Hawaii state T21 law | Average monthly cigarette sales in Hawaii | N/A | Average monthly cigarette sales in rest of US (excl. Alaska and California) | Change in average monthly unit sales: Hawaii -4.4%, USA -10.6% | Not tested | Serious |
| **Glover-Kudon 2021**[32] | Longitudinal cigarette sales data | 2012 - 2017 | California state T21 Law | Average monthly cigarette sales in California | N/A | Average monthly cigarette sales in rest of US (excl. Alaska and Hawaii) | Change in average monthly unit sales: California -11.7%, USA -10.6% | NA* | Serious |
| **Schneider 2016**[33] | Repeated cross-sectional | 2006-2012 | Needham, Massachusetts T21 | 9-12 graders living in Needham, Massachusetts (16 385 to 17 | U18 | Non-Needham residents in 16 surrounding areas (16 385 to 17 | Current cigarette smoking: reduction from 12.9% to 6.7% (interv.) 14.8% to 12.0% | Favours intervention | Critical |
|  |  |  | law | 089 per survey, four surveys)* |  | 089 per survey, four surveys)* | (control); p<0.01^ |  |  |
| **Schiff 2021**[34] | Cohort study | 2015 - 2017 | California state T21 Law | Southern California residents aged 19-20 post-policy (n=1609) | 18+ | Southern California residents aged 18-19 pre-policy (n=1502) | Past 30-day use, cigarette: Pre T21 = 150 (9.6%) Post T-21 = 164 (11.1%) | Not tested | Critical |
| **Hawkins 2022**[35] | Repeated cross-sectional | 2011 - 2017 | County laws in Massachusetts | Massachusetts resident in county with T21 (n = 9,988*) | U18 | Massachusetts resident in county without T21 (n = 9,988*) | Current cigarette smoking: Inflation model: 0.12 (-1.34 to 0.11) | Not significant | Moderate |
| **Liber 2022**[36] | Longitudinal cigarette sales data | 2015 - 2019 | T21 laws covering 60% of designated market areas | Nielsen Designated Market Areas with at least 60% coverage of T21 laws | N/A | Nielsen Designated Market Areas with less than 60% coverage of T21 laws | Diff-in-diff: Disproportionately young cigarette brand sales: -0.000156 (p = <0.001)^ | Favours intervention | Moderate |
| **Ali 2020**[37] | Longitudinal cigarette sales data | 2014 - 2018 | California state T21 law | California residents | N/A | Western states | Absolute adjusted risk difference of cigarette sales: -9.41 (-15.52, -3.30) ^ | Favours intervention | Serious |
| **Ali 2020**[37] | Longitudinal cigarette sales data | 2014 - 2018 | Hawaii state T21 law | Hawaii residents | N/A | Western states | Absolute adjusted risk difference of cigarette sales: **-0.57 (-0.83, -0.30)^** | Favours intervention | Serious |
| **Patel 2023**[38] | Cohort study | 2014 - 2019 | "Dose" of local, city or state T21 law | US residents exposed to T21 aged between 15-21 (n = 13,990*) | N/A | US residents not exposed to T21 aged between 15-21 (n = 13,990*) | Current cigarette use - model 1: AOR 0.90 (CI 0.72 - 1.14) | Not significant | Moderate |
| **Wilhelm 2021**[39] | Repeated | 2016 and 2019 | Minnesota city or | 8/9 graders living in Minnesota | U18 | 8/9 graders not living in | Current cigarette use, 8/9 grade: AOR 0.81 | Favours | Serious |
|  | cross-sectional |  | county T21 law | T21 area (210,177) |  | Minnesota T21 area (n = 210,177*) | (0.67, 0.99)^ | intervention |  |
| **Wilhelm 2021**[39] | Repeated cross-sectional | 2016 and 2019 | Minnesota city or county T21 law | 11 graders living in Minnesota T21 area (210,177*) | U18 | 11 graders not living in Minnesota T21 area (210,177*) | Cigarettes, 11 grade: AOR 1.2 (0.97, 1.48) | Not significant | Serious |
| **Yan 2014**[40] | Repeated cross-sectional | 1992 - 2002 | Pennsylvania state law | Women aged 20 - 20.99 years in Pennsylvania (n =60,710*) | 18+ | Pregnant women aged 21 - 22 years in Pennsylvania (n = 60,710*) | Prenatal smoking: 0.013 (0.010) | Not significant | Moderate |
| **Trapl 2022**[41] | Repeated cross-sectional | 2013 - 2019 | Cleveland City, OH T21 law | 9-12 graders living in Cleveland City (n=7,064) | U18 | 9-12 graders living in Cleveland suburbs (n=5,552) | Adjusted current cigarette smoking: ($\beta$ = 0.04 [SE, 0.07]; P=.56) | Not significant | Moderate |

\*Intervention group and compactor population size not reported separately (AOR = adjusted odds ratio, APR = adjusted prevalence ratio, ARR = adjusted risk ratio) ^ = statistically significant

The detailed results of the ROBINS-I assessments are set out in Supplementary File 4. These assessments relate specifically to single effect estimates, not entire studies. Estimates are very unlikely to be graded at a lower risk of bias than moderate using the ROBINS-I tool, given residual confounding introduced in non-randomised studies.[21] 10 estimates were judged to be of overall moderate risk of bias, 11 of serious risk of bias, three of critical risk of bias and two provided insufficient information for a judgement to be made. 10 estimates found an statistically significant association favouring T21, 12 estimates were not significant, one estimate favoured the control (MLSA of tobacco remaining at 18) and 3 estimates did not assess statistical significance (Figure 2).

**Figure 2:**
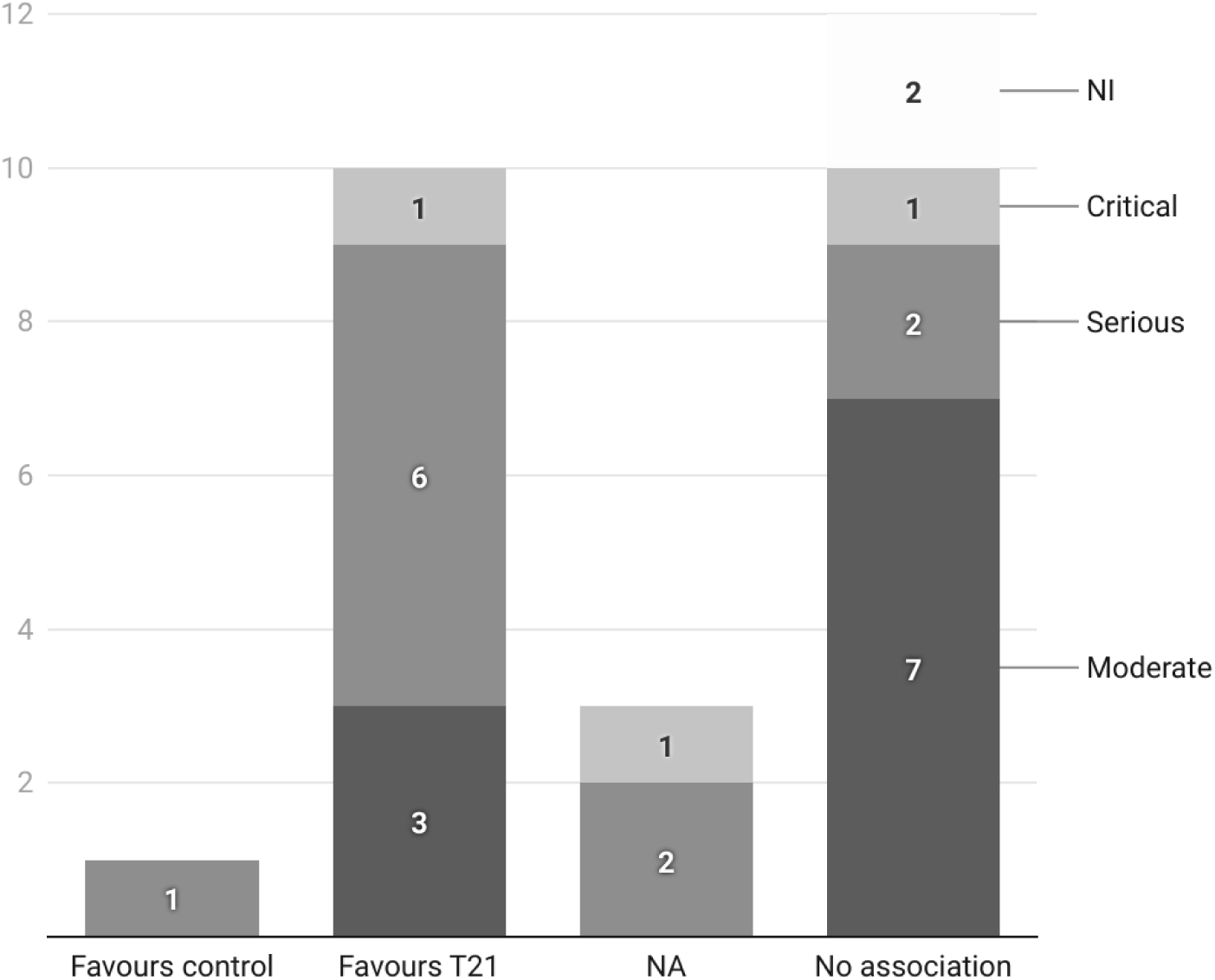
Direction of statistically significant association by risk of bias estimates

21 estimates were based on self-reported current smoking, of which 15 were based on repeated cross-sectional studies, three on one-off cross-sectional studies, and three were based on cohort studies. Five of the estimates were based on cigarette sales data over time.

### Age differentiation

We originally planned to conduct sensitivity analyses by age (11-17 and 18-20). Narratively, we found some evidence that there is a differential association of the T21 policy by age. Within estimates predominantly considering those aged 11-17, two estimates at moderate risk of bias found a significant association with T21 and reduced current cigarette smoking in older school-age groups, but not in younger school age-groups[29,35] and one study at critical risk of bias found stronger associations in older age groups.[33] In contrast, one study at serious risk of bias found a significant association in younger school-age groups, but not older school age group.[39] Analyses primarily considering those aged 18-20 were more likely to find a significant reduction in cigarette smoking linked with T21 than studies of younger age groups.

### Single law and multiple law evaluation

Analyses evaluating the impact of a single local, city or state law were less likely to report a positive association between T21 and reduced current smoking rates, with 13 of 17 analyses at serious, critical or NI risk of bias. This includes the study of New York’s T21 law, which conducted the only analysis to find an association between increased smoking rates and T21.[24] However, analyses that evaluated T21 across multiple geographies found significant associations between reduced current cigarette smoking and T21 on six of nine occasions, with all analyses at moderate or serious risk of bias.[23,26,29,36,38]

### Consideration of health inequalities

Seven studies considered differential associations of T21 by ethnicity or race. Studies defined ethnicity and race differently, making synthesis difficult; most notably, five analyses found a more pronounced association with T21 in Hispanic/Latinx groups than in White groups [25,27,29,41] with two analyses finding the opposite.[33,40]

Two studies at moderate risk of bias considered the differential associations of T21 by education.[29,40] Both found that T21 was associated more strongly with reduced cigarette smoking than for those with lesser parental or personal education.

One study at moderate risk of bias considered differential associations of T21 on current smoking by sexual minority and non-sexual minority and found little difference between groups.[25]

## DISCUSSION

To our knowledge, this is the first systematic review assessing the impact of laws raising the MLSA to 20 or above on smoking rates. Studies displayed a high level of heterogeneity in analytical approaches, population groups and the types of laws enforced. All studies were conducted in the US.

The totality of the evidence suggests that T21 policies are effective in reducing cigarette smoking, although the context and type of policy implementation is likely to be important. This is supplemented by two recent studies, not included in the systematic review as they were institutional working papers that were not peer reviewed. One paper found T21 was associated with a decline in smoking participation in both 16-17 year olds and 18-20 year olds[42] and one paper found a reduction in cigarette use for those in the 12^th^ grade, and in cigarette sales for counties with higher shares of under 21s.[43] Another study, not included in this systematic review, because it did not included data on smoking rates or cigarette sales, found lower smoking *intentions* amongst those with knowledge of T21 laws, [44] and a separate study found that the proportion of youth who perceived easy access to cigarettes significantly decreased following the federal T21 law.[45]

There were no studies at low risk of bias, which was expected given that the ROBINS-I assessment tool is extremely unlikely to assess non-randomised studies to be at low risk of bias. There were 21 separate analyses at moderate or serious risk of bias, a relatively rich source of evidence for a single tobacco control policy.

Age appeared to mediate the association with T21. Generally, the older the participant group, the more likely T21 was to be associated with reduced current cigarette smoking. The impact of T21 on the ability of 18-20 year olds and older teens to purchase tobacco may be more immediate. [44] For younger groups, more time might be required for T21 policies to change smoking habits, as it is more reliant on disrupting the supply of tobacco from older groups and wider changing social norms.[32]

We found that studies which included multiple geographical areas with T21 laws were more likely to find an association than those that evaluated a single area. Many of the early states and areas that introduced T21 and subsequently evaluated in isolation were already leaders in tobacco control, with relatively low prevalence rates amongst young people, and thus reducing cigarette smoking further is challenging. Many of the studies that focused on multiple areas included the impact of laws in parts of the country which had higher smoking prevalence [26–29,36,38] and thus the scope for reduction in current smoking may have been greater.

There were encouraging findings relating to T21’s potential for reducing racial and educational inequality in smoking, important given the extremely unequal impact of tobacco control on health.[1,46,47] Studies appeared to show that T21 was more strongly associated with reduced smoking status in Hispanic/Latinx and White groups, important as this group are less likely to use pharmacotherapy to make quit attempts once established smokers.[48] The policy was also shown to have the potential to reduce disparities in smoking rates across education level in two studies at moderate risk of bias, despite evidence there is a lower likelihood of ID checks in poorer areas. [49]

It is important to note large that in the US, rapid and significant increases in e-cigarette use took place during the duration of many of the included studies, along with falls in smoking rates. Only 1.5% of those in grades 9-12 were current cigarette users in 2021.[50] The literature on the association of T21 on e-cigarettes is beyond the scope of this review, but appears to be mixed. It is possible that this rise affected the impact of T21 laws, although some analyses controlled for e-cigarette use.[25,39] The relationship between MLSAs and e-cigarette use needs careful consideration given global increases in e-cigarette use in younger age groups.

T21 is not the only policy option for countries considering policies based on age of sale. The smoke-free or tobacco-free generation approach, in which the MLSA is effectively raised by one year every year, is a prominent alternative. New Zealand[17] and the UK [18] have started the process of implementing smoke-free generation laws and there are published modelling studies to support its implementation in various settings [51,52]

### Limitations

We were not able to conduct meta-analysis which would have provided more robust measures of association and uncertainty. However, the narrative synthesis that has been conducted is important to understanding the contexts in which Tobacco 21 is likely to have greater impact.

It was not clear from most studies how roll-your-own tobacco was categorised, although survey methodologies suggested this would have largely been included under cigarette smoking. This review does not consider e-cigarettes, cigars, or smokeless tobacco products. This may have prevented useful further context on product type; for example, Trapl et al’s paper found a significant reduction in cigar use in Cleveland compared to surrounding areas, though not cigarettes.[41] However, given a proliferation of estimates across product types, a focus on cigarette smoking enabled systematic, transparent approach to synthesising data for the outcome of greatest global relevance, given cigarette smoking is most common form of tobacco use in this age group.[1]

Importantly, all studies were conducted in the US. Evaluations of MLSAs above 20 in the several other countries which have implemented them will be critical to inform wider global tobacco control policymaking.

We did not look in-depth at the design and enforcement of laws and how this affected estimates, particularly important for MLSAs.[53] For example, California’s law exempted those serving in the military,[54] and some studies found that T21 implementation was accompanied by a lack of retailer monitoring, [24,55] an important component of ensuring compliance with new MLSAs.

## CONCLUSIONS

This review demonstrates that there is evidence that T21 policies are associated with reducing cigarette smoking and cigarette sales in those aged 11-20. T21 may be more likely to achieve its policy goals when implemented in areas with higher current cigarette smoking rates. There may be stronger associations with reduced smoking in older groups, Hispanic/Latinx groups and those with lower educational status, signifying a possible role in reducing health inequalities. Countries and regions with lower MLSAs should consider raising the age of sale of tobacco as part of a broader tobacco control strategy, paying careful attention to design, implementation, and enforcement of new laws.

## DECLARATIONS

The authors have no conflicts of interest to declare.

Contributions: ND: conceptualisation; study registration; data extraction; investigation; writing initial draft; review and editing. IB; study registration; data extraction; investigation; review and editing. SM: data extraction; investigation; review and editing. RM: conceptualisation; study registration; data extraction; investigation; review and editing. All authors contributed to the data interpretation, writing and revisions of the report and have full access to all data in the study.

Funding: This work was partly supported by NIHR award 302872.

## Supporting information

SuppFile 1_PRISMA checklist

SuppFile2_search strategy

SuppFile3_data extraction

SuppFile4_full RoB

## Data Availability

All data produced in the present study are available in the supplementary materials.

