## Supplementary material for "What is the relationship between raising the minimum legal sales age of tobacco above 20 and cigarette smoking? A systematic review": SuppFile2_search strategy

**Tobacco 20+ systematic review search strategy**

**Search strategy for MEDLINE through PubMed**

**(**Search: **((1. Tobacco OR exp. smoking OR cigarette* OR cigar* OR smok* OR nicotine*) AND (2. "age of sale" OR "age-of-sale" OR "sale age" OR "MLA" OR "MLSA" OR "minimum legal age" OR "minimum age" OR "age of legal access" OR "sale age" OR "age of purchase" OR "purchase age" OR "minimum purchas*" OR "legal minimum age" OR "age policy" OR "T21" OR "Tobacco 21" OR "MLA" OR "MLSA" OR "legal age" OR "age restriction")) AND (("2015/01/01"[Date - Publication] : "3000"[Date - Publication])) Filters: from 2015 - 2022**

**Search strategy for EMBASE through OVID**

**(**Tobacco or smoking or cigarette* or cigar* or smok* or nicotin*).mp. [mp=ti, ab, hw, tn, ot, dm, mf, dv, kf, fx, dq, tc, id, tm]

1. age of sale or age-of-sale or sale age or MLA or MLSA or minimum legal age or minimum age or age of legal access or sale age or age of purchase or purchase age or minimum purchas* or legal minimum age or age policy or T21 or Tobacco 21 or MLA or MLSA or legal age or age restriction
2. limit 2 to yr=”2015-Current”
3. 1 and 2 and 3

**Search strategy for PyschInfo (through OVID)**

1. (Tobacco or smoking or cigarette* or cigar* or smok* or nicotin*).mp. [mp=ti, ab, hw, tn, ot, dm, mf, dv, kf, fx, dq, tc, id, tm]
2. age of sale or age-of-sale or sale age or MLA or MLSA or minimum legal age or minimum age or age of legal access or sale age or age of purchase or purchase age or minimum purchas* or legal minimum age or age policy or T21 or Tobacco 21 or MLA or MLSA or legal age or age restriction
3. limit 2 to yr=”2015-Current”
4. 1 and 2 and 3

**Search strategy for ProQuest Public Health Database and Dissertations and Theses**

(Tobacco OR exp. smoking OR cigarette* OR cigar* OR smok* OR nicotin*) AND ("age of sale" OR "age-of-sale" OR "sale age" OR "MLA" OR "MLSA" OR "minimum legal age" OR "minimum age" OR "age of legal access" OR "sale age" OR "age of purchase" OR "purchase age" OR "minimum purchas*" OR "legal minimum age" OR "age policy" OR "T21" OR "Tobacco 21" OR "MLA" OR "MLSA" OR "legal age" OR "age restriction") Date: after 01 January 2015

**Search strategy for CINHL through Ebscohost**

| ( 1. Tobacco OR exp. smoking OR cigarette* OR cigar* OR smok* OR nicotin* ) AND ( 2. “age of sale” OR “age-of-sale” OR “sale age” OR “MLA” OR “MLSA” OR “minimum legal age” OR “minimum age” OR “age of legal access” OR “sale age” OR “age of purchase” OR “purchase age” OR “minimum purchas*” OR “legal minimum age” OR “age policy” OR “T21” OR “Tobacco 21” OR “MLA” OR “MLSA” OR “legal age” OR “age restriction ) |
| --- |

Limiters - Published Date: 20150101-20231231
