## Supplementary material for "What is the relationship between raising the minimum legal sales age of tobacco above 20 and cigarette smoking? A systematic review": SuppFile4_full RoB

**Supplementary material x: full risk of bias assessment**

| **Paper** | **Intervention location** | **Numerical result being assessed** | **Confounding** | **Participant selection** | **Classification of interventions** | **Deviation from intended intervention** | **Missing data** | **Measurement of outcomes** | **Selection of reported result** | **Overall risk of bias** |
| --- | --- | --- | --- | --- | --- | --- | --- | --- | --- | --- |
| **Yan 2014** | Pennsylvania | Prenatal smoking: 0.013 (0.010) (Table 2, column 8) | Moderate | Low | Low | Moderate | Low | Moderate | Low | **Moderate** |
| **Ali 2022** | Hawaii | Versus intervention states: -0.57 (-0.83 to -0.30) (Table 2) | Serious | Low | Low | Low | Low | Low | Low | **Serious** |
| **Ali 2022** | California | Versus intervention states: -9.41 (-15.52 to -3.30) (Table 2) | Serious | Low | Low | Low | Low | Low | Low | **Serious** |
| **Agaku 2022 - YRBS** | States implementing T21 | OR 0.70 (CI 0.52-0.93) (Table 4) | Serious | Low | Moderate | Low | NI | Low | Low | **Serious** |
| **Agaku 2022 - BRFSS** | States implementing T21 | OR 0.58 (0.39 - 0.74) (Table 3) | Serious | Serious | Moderate | Low | NI | Low | Low | **Serious** |
| **Colston 2022** | Covered by local, county or state law | Smoking participation for 8 grader: ARR 0.91 (0.69, 1.20) (with multiple imputation) (Table 2) | Low | Low | Low | Low | Low | Low | Moderate | **Moderate** |
| **Colston 2022** | Covered by local, county or state law | Smoking participation for 10 grader = ARR 0.91 (0.69, 1.20) (with multiple imputation) (Table 2) | Low | Low | Low | Low | Low | Low | Moderate | **Moderate** |
| **Colston 2022** | Covered by local, county or state law | Smoking participation for 12 grader = ARR 0.74 (0.60,0.91) (with multiple imputation) (Table 2) | Low | Low | Low | Low | Low | Low | Moderate | **Moderate** |
| **Friedman 2019** | Various localities | OR 0.61 (95% CI 0.42, 0.89) Table 3, column 2, row 3 | Serious | Serious | Low | Low | Low | Low | Low | **Serious** |
| **Friedman 2020** | Various localities | Tobacco-21 policy covering entire MMS = -0.0306 reduction in 18-20 year olds (CI -0.0548 to -0.0063) - difference in difference, 18-20 (para 3 of results, top-left cell Table 2) | Low | Low | Moderate | Low | Low | Low | Low | **Moderate** |
| **Garcia-Ramirez 2022** | California | OR 0.98 (0.94, 1.03) (Table 1, Model 1, T21) | Moderate | Low | Low | Low | Moderate | Low | Low | **Moderate** |
| **Glover-Kudon 2021** | California | California −11.7%**, USA −10.6%** (Table 1) | Moderate | Serious | Low | Low | Low | Low | Low | **Serious** |
| **Glover-Kudon 2021** | Hawaii | Hawaii −4.4%**, USA −10.6%** (Table 1) | Moderate | Serious | Low | Low | Low | Low | Low | **Serious** |
| **Grube 2021** | California | Past 30-day cigarette smoking: OR 0.99 (0.97 - 1.01) (Table 1, Column 3) | Moderate | Low | Low | Low | NI | Low | Low | **Moderate** |
| **Hawkins 2022** | Localities in Massachusetts | 0.12 (CI = -1.34 to 0.11) (table 1, row 2, inflation model) | Moderate | Low | Low | Low | Moderate | Low | Low | **Moderate** |
| **Liber 2022** | Various | Diff-in-diff change in disproportionately young brands : −0.00156 (p = <0.001) (Table 3) | Moderate | Moderate | Low | Low | Low | Low | Low | **Moderate** |
| **Macinko 2018** | New York (vs New York state) | APR 1.25 (0.88, 1.76) (Table 3, column 2) | Low | Low | Serious | Low | NI | Low | Low | **Serious** |
| **Macinko 2018** | New York (vs FL counties) | APR 1.40 (1.10, 1.80) (Table 3, column 2) | Moderate | Low | Low | Low | NI | Low | Low | **NI** |
| **Roberts 202** |  | 1st years 2016 (comparator) - 6.6%. 1st year 2018 (intervention) - 4.1%. (Table 2) | Critical | Serious | Low | Low | Moderate | Low | Low | **Critical** |
| **Patel 2022** | Various | 0.90 (CI 0.72,1.14) Table 3, Model 1 | Low | Moderate | Low | Low | Moderate | Low | Low | **Moderate** |
| **Schneider 2015** | Needham, Massachusetts | -1.08, p<0.001 (Table 1, column 1) | Serious | Serious | NI | Low | Moderate | Low | Serious | **Serious** |
| **Schiff 2021** | California | Pre T21 = 150 (9.6%) Post T-21 = 164 (11.1%) (Table 1) | Critical | Low | Low | Low | Serious | Low | Low | **Critical** |
| **Trapl 202** | City of Cleveland | (β = 0.04 [SE, 0.07]; P=.56) (Table 3, row 1) | Moderate | Low | Moderate | Low | Moderate | Low | Low | **Moderate** |
| **Wilhelm 2022** | Minnesota T21 area | Current cigarette use, 8/9 grade: **AOR** **0.81 (0.67, 0.99)** (Table 2) | Serious | Moderate | Low | Low | Serious | Low | Moderate | **Serious** |
| **Wilhelm 2022** | Minnesota T21 area | Cigarette use, 11 grade: 1.2 (0.97, 1.48) (Table 2) | Serious | Moderate | Low | Low | Serious | Low | Moderate | **Serious** |
